# Pathogenic mitochondrial genome variation, heteroplasmy thresholding and mitochondrial constraint measures in a healthy older cohort

**DOI:** 10.64898/2026.06.24.26356403

**Authors:** Eloise Watson, Gordon Qian, Shyamsundar Ravishankar, Matthew Hobbs, Joseph Copty, Chenglong Yu, Sarah Kummerfeld, Christina Liang, Paul Lacaze, Ryan L. Davis, Carolyn M. Sue

## Abstract

Mitochondrial diseases (MDs) are clinically heterogeneous rendering ascertainment challenging. Estimates of pathogenic mitochondrial DNA (mtDNA) variants in the population range from 1 in 200 to 1 in 4,000 individuals. Inclusion of mtDNA sequencing in genomic databases facilitates comprehensive estimation of mtDNA variation. However, interpretation of low heteroplasmy variation is complex, due in part to misalignment of nuclear mitochondrial DNA transcripts (NUMTs), whilst conservative heteroplasmy thresholds likely omit relevant variation. Cumulative burden of mtDNA variation contributes to aging and neurodegeneration, and recent characterisation of mitochondrial genome constraint allows quantitation of this burden.

We analysed whole genome sequencing of blood DNA from 3,500 healthy older individuals in the Medical Genome Reference Bank using *mity*, considering pathogenic mtDNA variants ≥1% heteroplasmy. We identified 34 distinct pathogenic mtDNA variants in 62 individuals, giving a combined population allele frequency of 1.77% (95% CI 1.36-2.27) or 1 in 56 individuals. We evaluated inclusion of false positive (FP) calls due to two common NUMTs, which accounted for up to 16% of variants. Increasing heteroplasmy thresholding to eliminate all NUMT-FPs also eliminated much of the total variation, including pathogenic variants. We propose a sample-specific, scaled heteroplasmy threshold to maximise variant retention and mitigate NUMT-FPs. Finally, we characterised measures of mitochondrial constraint in this healthy older cohort, observing an association between variant burden and summed constraint, whilst mean constraint was higher in pathogenic variant carriers.

These findings suggest pathogenic mtDNA variation is more common in the population than is currently appreciated. Findings are comparable to larger genomic databases when heteroplasmy thresholding is adjusted, and support earlier population-based estimates. Incorporation of low heteroplasmy variation is relevant, but interpretation is nuanced, and optimising variant retention requires consideration of NUMT-FP rates.

## Introduction

Mitochondrial diseases (MDs) are caused by mutations in either the nuclear (nDNA) or mitochondrial (mtDNA) genome.[1] They exhibit considerable genetic and clinical variability, and as diagnostic capabilities advance, the range of recognised MDs continues to expand.[2–5] Although MDs are considered the most common form of inherited metabolic disease, [1] estimates of pathogenic mtDNA variation, MD risk and disease prevalence vary widely across studies, predominantly due to differences in research methodologies [6–23]. Estimates from clinical disease cohorts suggest 1 in 4,300 individuals harbour pathogenic MD variants, whilst 1 in 8,000 adults and approximately 1 in 5,000 individuals manifest disease, [8, 9, 11–13, 16, 19–21] although acknowledge ascertainment is likely incomplete. Conversely, studies utilising population-based approaches indicate that the number of individuals harbouring pathogenic mtDNA variants, and who are therefore potentially at risk of MD, may be as high as 1 in 200,[14, 15, 17, 18, 24] although examined only one or a small number of mtDNA variants. The limited phenotypic data associated with these population-based studies [14, 18, 24] suggest that there may be minor or subclinical disease manifestations that are not recognised as MD in practice. The substantial disparity between clinical cohort and population-based estimates along methodological lines reveals an important persisting gap in our understanding of mitochondrial variation in the population. This uncertainty extends to our understanding of the development and recognition of disease, which may have important implications for clinical practice and healthcare resource allocation.[25, 26]

Advances in sequencing technology, including bigenomic sequencing obtained simultaneously from whole genome sequencing (WGS), mean population-based approaches at scale are now feasible. Recent incorporation of mitochondrial variant frequencies in population databases, such as the Genome Aggregation Database (gnomAD),[27] Helix Mitochondrial Database, HelixMTdb [28] and UK Biobank [29], offers a population-wide scope to evaluate pathogenic mtDNA variation. However, sequencing of the small, circular mtDNA molecule presents unique considerations, including differentiation of nuclear mitochondrial DNA transcripts (NUMTs) from genuine mtDNA variation [30–32], and unique characteristics of mitochondrial genetics (e.g., heteroplasmy, tissue-dependent mutation load and the threshold effect),[33, 34] make interpretation of sequencing output complex.

Bioinformatic tools are continuously evolving to confidently identify mtDNA variants down to and below 1% heteroplasmy. However, clinical reporting typically implements heteroplasmy thresholds of 5-10% and open genomic databases such as gnomAD have prioritised conservative reporting with a high minimum heteroplasmy cut-off of 10%. Whilst this minimises NUMT contamination and risk of false positive variant calls, it likely underestimates genuine mtDNA variation, including pathogenic variants. Cannon and colleagues reported mtDNA variation in the UK Biobank down to 3% heteroplasmy with correspondingly higher rates of relevant variant identification. Bolze and colleagues utilised exome-based mtDNA sequencing on buccal cell samples, reporting heteroplasmy down to 0.7%, albeit with low average mtDNA coverage (182×) and limited haplogroup diversity. These considerations are particularly relevant with regard to mtDNA sequencing from blood, given variants occur at lower heteroplasmy due to high turnover and negative selection, such that very low blood heteroplasmy levels (e.g. <1%) have been identified in symptomatic individuals with m.3243A>G associated mtDNA disease.[35] Therefore, considering low heteroplasmy variation in the population may be of direct clinical relevance in an era of increasing reliance on comprehensive genome sequencing in clinical practice.

Expanding our understanding of pathogenic mtDNA variation across the heteroplasmy spectrum in the healthy population is essential for accurately contextualising variants identified in clinical sequencing results.[36, 37] Crucially, this effort must be paired with detailed phenotypic information allowing robust insight into variant penetrance, including atypical and oligosymptomatic presentations, to properly inform associated risks and genetic counselling for carriers. Collectively, such data could provide deep insights into the mechanisms influencing disease expression, as well as quantifying the potential burden of sub-clinical cases that present to medical specialties with isolated symptoms.

Understanding mtDNA variation beyond specific pathogenic variants and MDs may also further understanding of aging and neurodegenerative processes. Mitochondrial dysfunction is implicated in these processes through various pathomechanisms,[38, 39] such as the accumulation of typically low heteroplasmy somatic variation observed with age.[40, 41] Recent work has quantified nucleotide constraint across the mitochondrial genome [42] offering a new avenue to explore the burden of mtDNA variation. Conceptually, constraint metrics may quantify aggregate mitochondrial genome quality. This may offer a biomarker, biological insights into mitochondrial dysfunction, aging and neurodegeneration, and deeper understanding of MD penetrance and expression.

To characterise and understand the disparity between population-based and disease cohort estimates of MD prevalence, we utilised the sensitive mtDNA variant detection tool, *mity.*[43] Specifically, this study evaluated the frequency of pathogenic mtDNA variation down to low heteroplasmy in individuals from the Medical Genome Reference Bank (MGRB), a bigenomic whole genome-sequenced biobank of healthy, older individuals.[40] The potential impact of both polymorphic NUMT contamination and heteroplasmy thresholding on mtDNA sequencing results and pathogenic mtDNA variant prevalence was thoroughly evaluated to optimise detection thresholds. We compared the MGRB with data from large, publicly accessible genomic cohorts, as well as prior population-based and clinical cohort estimates, to place our findings in broader context for further research of MD, aging and neurological disorders, and for clinical practice. Finally, we evaluated mtDNA nucleotide constraint as a possible benchmark for studying mitochondrial genome variation in aging and other complex conditions.

## Methods

### Cohort

We utilised MGRB WGS data from blood DNA of 3,512 healthy older (64-95 years) Australians with no prior history of diagnosed cancer, cardiovascular or major neurological disorders [40] recruited from the *ASPREE* [44] and *45 and Up* (*45nUp*) [45] studies. Cohort rationale, participant selection, ethical governance and the bigenomic sequencing pipeline has been detailed previously [40, 46], with subsequent incorporation of additional genomes totalling 3,512 individual genomic samples. Comparative data from MitoMap, gnomAD (with and without low heteroplasmy filter) and HelixMTdb were drawn directly from the online databases (mitomap.org/MITOMAP, [47] gnomad.broadinstitute.org/ [27] and helix.com/mitochondrial-variant-database [28] respectively). Comparative data from UK Biobank was drawn from the published work by Cannon *et al*.[29]

### mtDNA variation, haplogroups, coverage and copy number

For this study, mtDNA variants were called using *mity* version 0.4, a sensitive mitochondrial variant analysis tool and developed specifically for mtDNA variant calling from WGS data.[43] Due to linearisation of the circular mitochondrial genome for alignment with the rCRS (GenBank Accession NC_012920), coverage in the hypervariable control region (m.16024-576) is artificially reduced, and therefore this region is not considered when using *mity*.[43] To balance clinical relevance of identified mtDNA variation with the spectrum of previously observed blood heteroplasmy of pathogenic mtDNA variants,[35] whilst minimising risk of misalignment with NUMTs, only pathogenic mtDNA variants with heteroplasmy of 1% or greater and at least 10 reads (*mity* Tier 1 variants) were considered in the analysis. Pathogenic mtDNA single nucleotide variants (SNVs) were defined using MitoMap’s list of confirmed pathogenic variants (mitomap.org/foswiki/bin/view/MITOMAP/ConfirmedMutations, [47] as at last update 01/03/2025). Carrier frequencies and 95% confidence intervals (CI) were calculated using the exact binomial method.

mtDNA haplogroups and haplotagging single nucleotide polymorphisms (SNPs) were determined using Haplogrep 3.2.1 [48, 49] and PhyloTree Build 17.2.[50] Haplogroup enrichment of pathogenic variants was calculated based on the relative proportion of haplogroups amongst individuals carrying a pathogenic variant compared to the proportional representation of haplogroups in the cohort.

mtDNA copy number (mtCN) was calculated according to Ding *et al*, 2015,[51] based on the ratio of mean nuclear to mean mitochondrial reads across each genome:

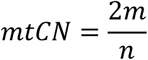

where *n* is mean nuclear coverage and *m* is mean mitochondrial coverage. Linear regression was used to evaluate the association of variant frequency and batch-adjusted mtCN with age and Welch two-sample *t*-testing was used to compare mtCN by sex. Kruskal-Wallis testing was used to evaluate the difference in mean proportions of variants in different heteroplasmy bins by age group, followed by pairwise Wilcoxon rank-sum tests between each age group pair.

### NUMT analysis and heteroplasmy thresholding

Evaluation of polymorphic NUMT inclusion in the MGRB *mity* dataset focused on 25 common NUMT false positive (NUMT-FP) variants from two polymorphic NUMT regions identified by Laricchia *et al.*.[27] numtA is an 871-bp insertion of Chr M: 12,361-13,277 into Chr 21:9,676,568, contributing 5 NUMT-FP variants, and numtB is a 536-bp insertion of Chr M:16093-59 into Chr 11: 49,862,017, contirbuting 20 NUMT-FP variants (**Suppl 1_NUMT FPs**). NUMTs preferentially involve non-coding mtDNA [30] and as numtA was localised in the hypervariable region, this region was retained for the purposes of this analysis. The 25 NUMT-FP variants were used to calculate a “floor” estimate of NUMT-FP inclusion in the MGRB *mity* dataset (numtA and numtB, as two confirmed exemplars of NUMTs, were used to represent the many other known polymorphic and unknown private NUMTs [30, 32, 52]). To compare theoretic versus observed NUMT-FP heteroplasmy, the theoretic variant allele fraction (VAF; in this context equivalent to heteroplasmy) at which a heterozygous NUMT-FP variant would be observed in individual WGS samples was approximated based on the ratio of nuclear to mtDNA coverage:

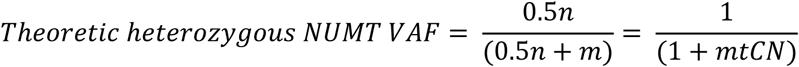

where *n* is mean nuclear coverage and *m* is mean mitochondrial coverage. For this component of the analysis, as well as incorporating the hypervariable region, the ≥1% heteroplasmy threshold in the MGRB *mity* dataset was removed, incorporating all variants with a minimum of 10 alternate reads (Tier 1 and 2 *mity* variants). A two-component finite mixture model (fleximix, an unsupervised clustering method) was fitted to the relationship between theoretical and observed NUMT-FP VAF. The fit lines were constrained to pass through the origin, and the resultant component slopes were compared to theoretical expectations for heterozygous (*y* = *x*) and homozygous (*y* = 2*x*) NUMT-FPs. This analysis informed a sample-specific heteroplasmy thresholding approach, based on theoretic NUMT allele frequency for the sample, which was applied to explore optimisation of total variant retention, pathogenic variant detection sensitivity, and NUMT-FP variant contamination. The effect of varied heteroplasmy thresholding on variant identification and prevalence estimates was then compared between the MGRB *mity* dataset and publicly available databases including gnomAD,[27] UK Biobank[29] and HelixMTdb.[28]

### Mitochondrial constraint measures

For evaluation of mitochondrial genome constraint, inclusion of somatic variation – typically at low heteroplasmy – is particularly relevant. Therefore, we included all variants in the dataset with heteroplasmy ≥1%, including those in the hypervariable control region. Following removal of sample haplotagging SNPs (as identified by Haplogrep v3.2.1[49]) from the data, we identified and removed common NUMT-FPs. Remaining single nucleotide variants (SNVs) were used to calculate constraint scores. mtDNA local constraint (MLC) scores for SNVs, drawn from Lake *et al*., 2024,[42] were applied to all SNVs identified in the MGRB dataset. MLC scores for each SNV in an individual sample were summed to give a summed constraint score (SCS) for each individual sample, and divided by the total number of contributing SNVs in the sample to give a mean constraint score (MCS) for each sample. Linear regression was used to evaluate associations between constraint measures, number of variants and age, and Welch’s two-sample *t*-test was used to compare differences in constraint measures between pathogenic variant carriers and non-carriers.

All analyses were performed using R studio version 4.0.5.

## Results

Average nuclear sequencing coverage was 38× (38.38×; standard deviation (sd) 3.34; range 26.85-46.93), whilst average coverage across the mitochondrial genome was greater than 3,300× (3,367.13×; sd 1681.15; range 469.42-14454.1). Sequencing depth was largely consistent across the mitochondrial genome, excepting the control region (m.16,024-576), owing to alignment of this circular genome contig to the linear rCRS, and the invariant ‘n’ spacer at m.3107. Average mtDNA copy number (mtCN) was 174.93 copies per sample (174.93; sd 83.96; range 28.26-694.42 copies; **Supplementary Figure 1**). There was an inverse association observed between mtCN and increasing age (decline of 0.94 copies per year; *p* = 0.000236) and a direct relationship between variant frequency and age (increase of 0.25 variants per year, *p* = 0.14×10^−11^) (**Supplementary Figure 2**), with no significant difference in these associations by cohort. Consistent with previous reports, [51] there was a slightly higher representative mean mtCN in females (177.8 vs 171.03; p = 0.016). Four samples with unexpectedly low nDNA coverage and therefore disproportionately high mtCN, as well as eight samples with unexpectedly high mtDNA depth (15,000×) and therefore mtCN (>750) were excluded. Of the 3,500 remaining samples, 2,789 derived from the ASPREE cohort (age ≥75) and 711 from the 45nUP cohort (age 64-95). The mean age of subjects was 78 years (78.07; sd 5.19; range 64-95 years) and a slight majority (58%) were female. Haplogroup distribution was consistent with the known European-predominant population structure of Australia [40], with haplogroup H most commonly observed, (*n* = 1550; 44.29%), followed by U (*n* = 479; 13.69%), J (*n* = 386; 11.03%), T (*n* = 378; 10.8%), K (*n* = 286; 8.17%), I (*n* = 114; 3.26%) and V (*n* = 109; 3.11%). The remaining 198 samples (5.66%) comprised a mixture of haplogroups (W (71; 2.03%), X (42; 1.2%), M (32; 0.91%), R (13; 0.37%), N (12; 0.34%), L (9; 0.26%), B (6; 0.17%), D (4; 0.11%), A, C and S (each 2, 0.06%) and F, G and Z (each 1; 0.03%).

Across the mitochondrial genome, variation was unsurprisingly common. Although variants were observed at all heteroplasmy levels, variants at the extremes of heteroplasmy were most common. Most variants were found at heteroplasmy below 1% (204,055/368,098; 55.43%) but due to a higher potential for sequencing artefact, these variants were not considered in analysis for pathogenic variants. Additional exclusion of variants with low total sequencing depths (<200) and those in the control region left a total of 93,316 variants for consideration (**Figure 1**). Of these, over half (52,960 variants; 56.75%) were homoplasmic or near homoplasmic (heteroplasmy ≥95%); the majority of which (49,688 variants; or 93.82% of near/homoplasmic variants and 53.25% of total variants) had heteroplasmy of 99% or more. Less than 2% of variants (1,132 variants; 1.29%) fell in the 10-95% heteroplasmy range, whilst variants in the 1-10% low heteroplasmy range accounted for 42% of variants (39,227 variants), of which the majority (32,382 variants; 82.55% of low heteroplasmy and 34.7% of total variants) were between 1 and 2% heteroplasmy and a further 4,319 variants (11.01% of low heteroplasmy variants and 4.63% of total variants) were at heteroplasmies between 2-3%.

**Figure 1.**
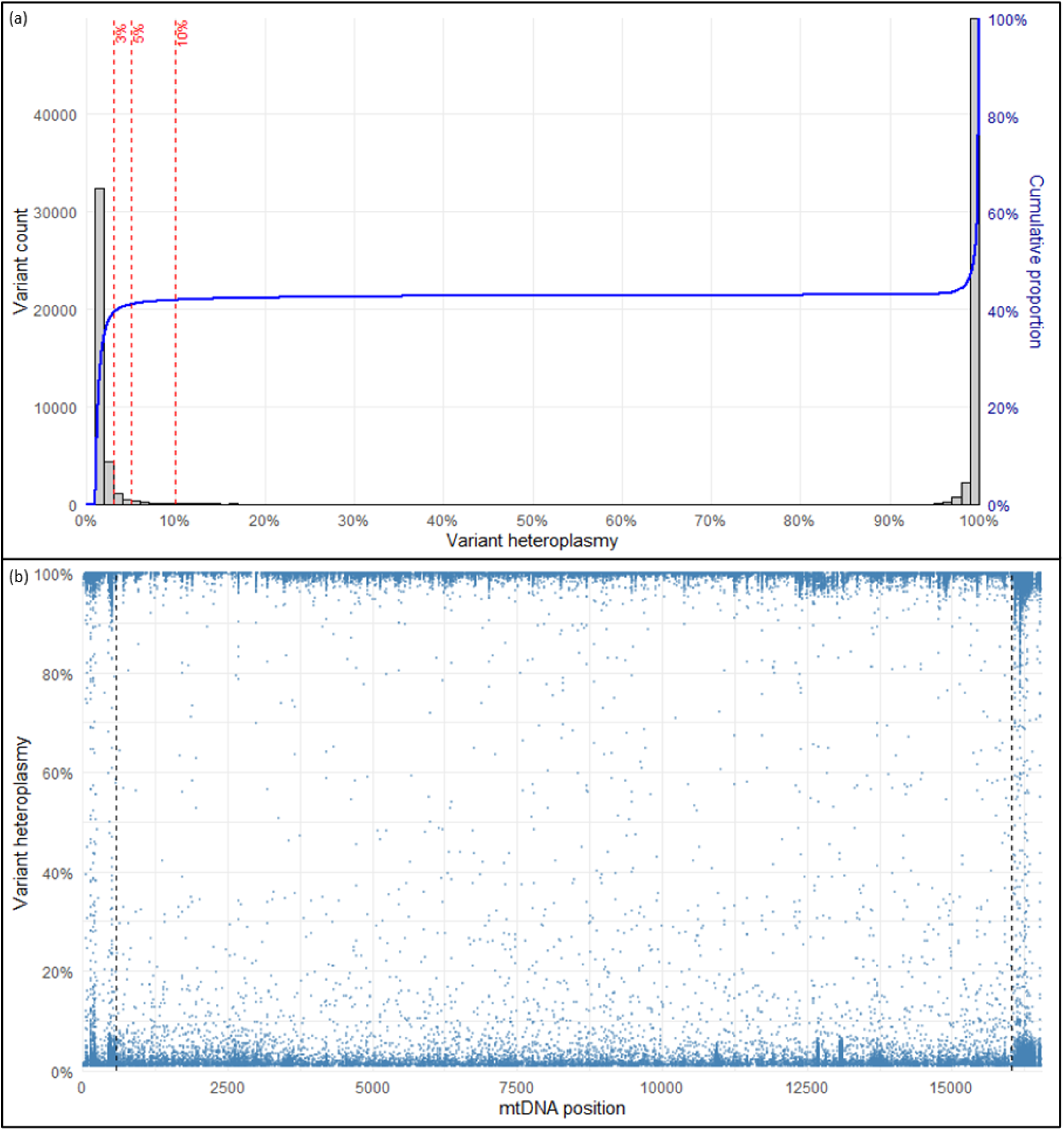
Distribution of mtDNA variation by heteroplasmy and variant heteroplasmy by base position in MGRB samples; **(a)** variant count (bars) and cumulative variant proportion (cumulative distribution function) by heteroplasmy for variants with heteroplasmy ≥1%, excluding the hypervariable region and variants with low read depth. Vertical red dashed lines indicate heteroplasmy thresholds of 3%, 5% and 10%. Distribution of variant heteroplasmy by base position is depicted in **(b)**, demonstrating clustering of variation towards very high and very low heteroplasmy. Black vertical dashed lines delimit the hypervariable control region.

Exclusive consideration of the 54,092 variants with heteroplasmy between 10 and 100% revealed proportions of heteroplasmic variants and homoplasmic or near homoplasmic variants, at 2.1% and 97.9% respectively. This was remarkably similar to proportions reported in the gnomAD cohort of 1.9 million variants, where 98% were homoplasmic or near homoplasmic and 2% were heteroplasmic.[27] The observed mean number of heteroplasmic variants per individual in this heteroplasmy range was also similar to that reported in the gnomAD database.[27] Considering all variants with heteroplasmy ≥1%, including homoplasmic variants, the average number of variants per individual was 26.66 variants (range 3-120), with a significantly higher relative proportion of low heteroplasmy variants in older individuals, and conversely a significantly lower relative proportion of high heteroplasmy variants (**Supplementary Figure 3**). This is consistent with the observation of increasing variation with age and indicative of accumulation of somatic variation, although the absolute difference was small compared to total variation.

### Population frequency of identified pathogenic variants

A total of 62 pathogenic variants with heteroplasmy of ≥1% (mean read depth 474 and median 62.5; range 18-3721) were identified in 62 individuals (no individuals carried more than one pathogenic variant) and represented 34 distinct pathogenic variants (**Table 1**). Combined population allele frequency of pathogenic mtDNA variants (heteroplasmy ≥1%) was therefore calculated to be 1.77% (95% CI 1.36-2.27), or 1 in 56 individuals. Nearly half of the pathogenic variants (*n* = 28; 45%) were present at heteroplasmy >2.5%, and almost one third (*n* = 19; 31%, or 1 in 184 individuals) had heteroplasmy of 5% or greater. Approximately one quarter (*n* = 15; 24%) of pathogenic variants met a more stringent cut-off (such as that implemented by gnomAD) of 10% heteroplasmy, indicating that one in 233 individuals in this cohort carried a pathogenic mtDNA variant meeting a stringent threshold of 10% heteroplasmy or higher. This accords closely with the findings of Laricchia *et al*., who found that 1 in 250 individuals carry a pathogenic variant with heteroplasmy of 10% or higher (**Table 2**).[27]

**Table 1:**
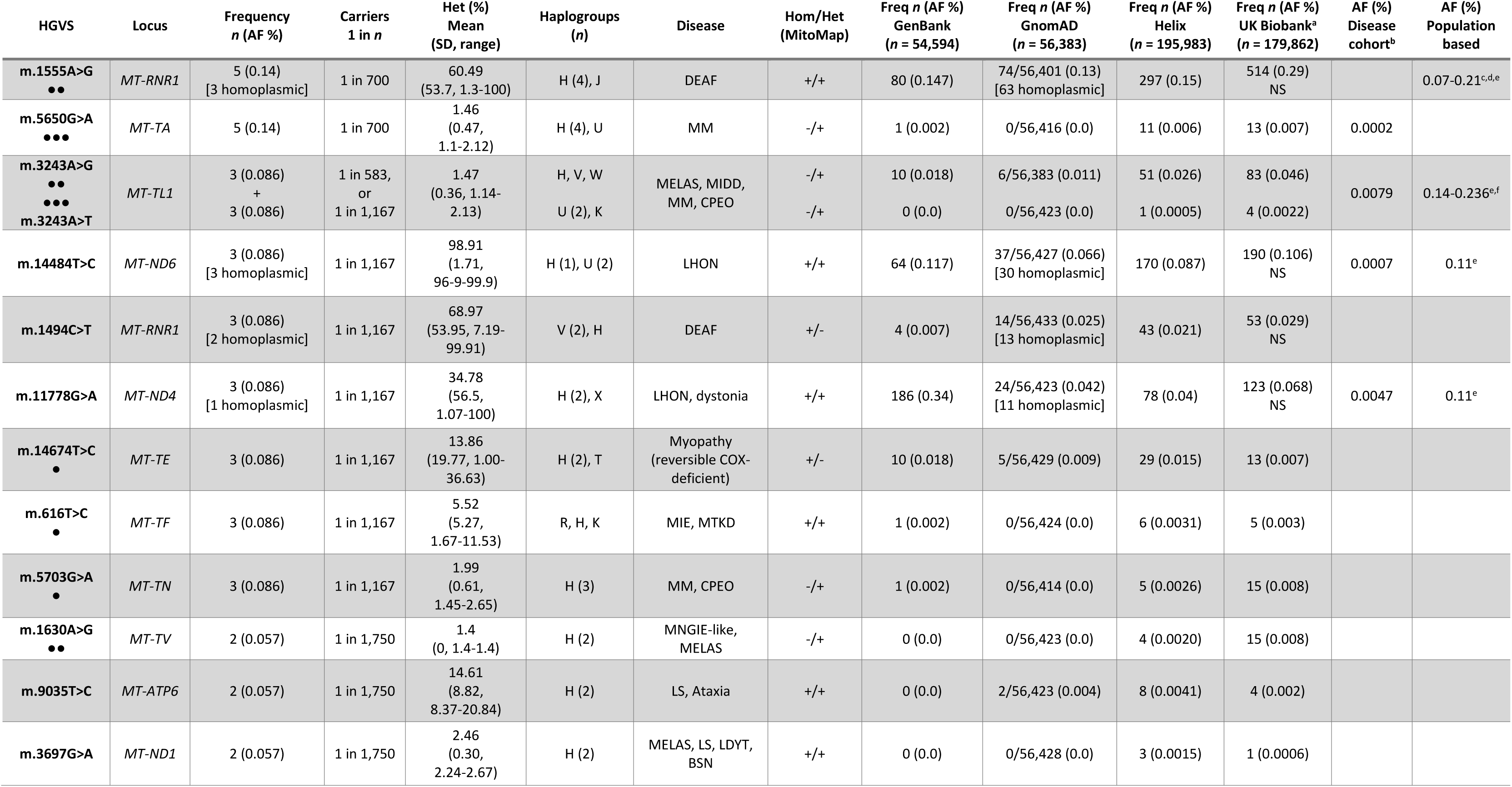
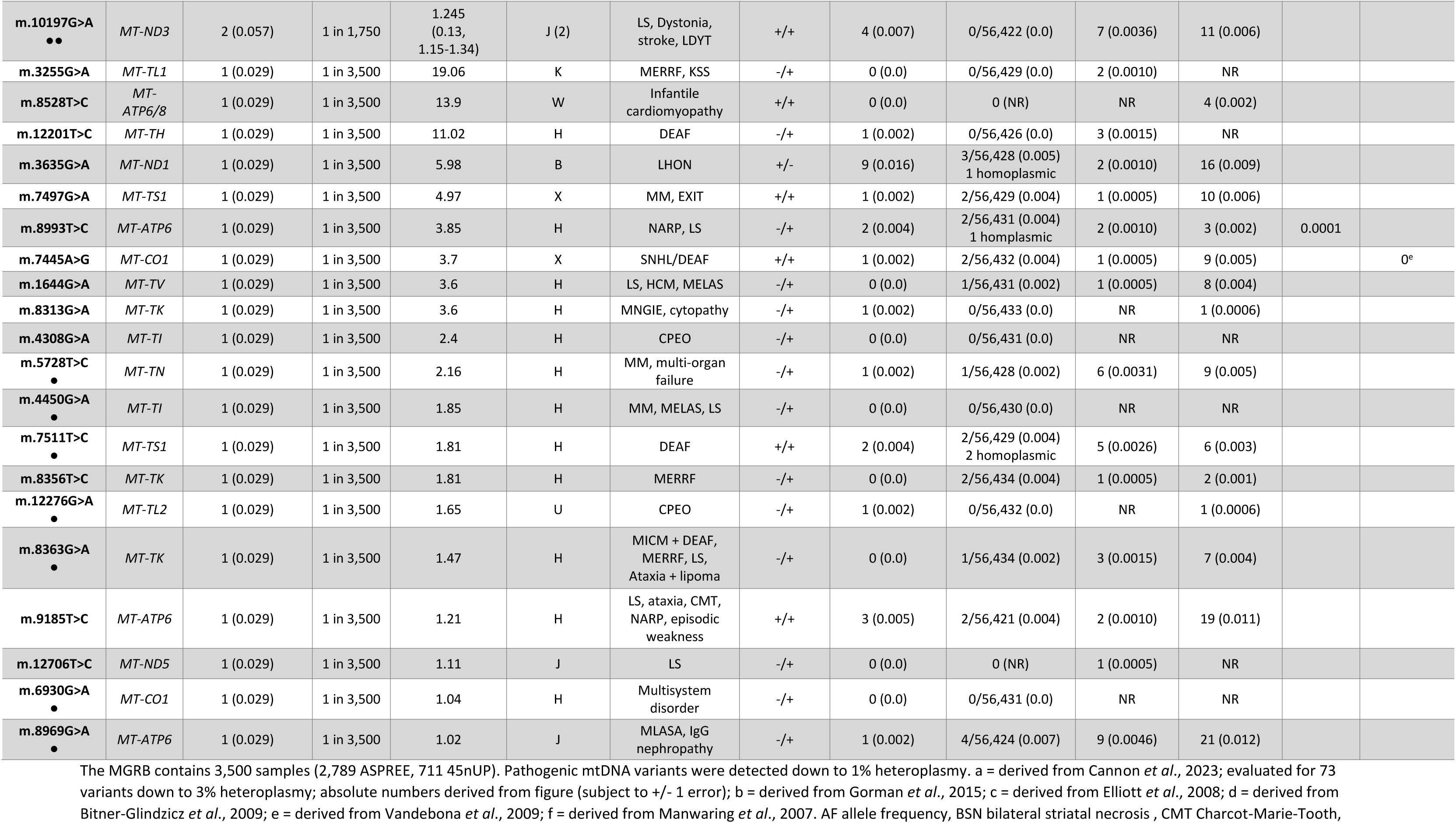
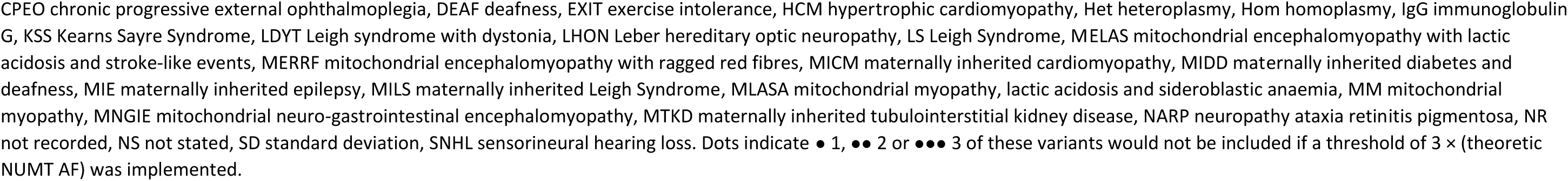
Pathogenic mtDNA variants identified in the MGRB compared to population databases, disease-cohort and population-based study estimates.

**Table 2.** Comparison of summed pathogenic mtDNA variant allele frequencies between different genomic databases applying varied heteroplasmy thresholds, with estimates from earlier disease-cohort based and population-based studies.

| For all MitoMap confirmed pathogenic mtDNA variants | Summed alleles | Total genomes | Summed AF (%) | 1 in *n* | *n* per 100,000 |
| --- | --- | --- | --- | --- | --- |
| **MGRB; heteroplasmy ≥1%** | 62 | 3,500 | 1.77 | 56 | 1,771 |
| **MGRB; sample-specific threshold: 2×theoretic NUMT AF** | 49 | 3,500 | 1.4 | 71 | 1,400 |
| **MGRB; sample-specific threshold: 3×theoretic NUMT AF** | 38 | 3,500 | 1.08 | 92 | 1,086 |
| **MGRB; 10% heteroplasmy threshold** | 15 | 3,500 | 0.43 | 233 | 428 |
| **gnomAD; incorporating low heteroplasmy ≥1%** | 657 | 56,434 | 1.16 | 86 | 1,164 |
| **gnomAD; default 10% heteroplasmy threshold** | 204 | 56,434 | 0.36 | 277 | 361 |
| **UK Biobank (Cannon, 2023); heteroplasmy ≥3%** | 1,295 | 179,862 | 0.72 | 139 | 720 |
| **HelixMTdb (WES)** | 846 | 196,554 | 0.43 | 232 | 430 |
| **Disease cohort-based: Gorman, 2015** | - | - | 0.020 | 4,902 | 20.4 |
| **Population-based: Elliott, 2008; 10 common mtDNA variants** | 15 | 3,168 | 0.54 | 211 | 540 |
| Manwaring, 2007; m.3243A>G \| Vandebona, 2009; m.1555A>G | 6 \| 7 | 2,954 \| 2,856 | 0.45 | 223 | 447 |

Except where specified, summed alleles reflect sum of all confirmed pathogenic variants (from MitoMap) in each database. The lower portion of the table contains indicative examples from earlier disease cohort-based, and population-based studies for comparison. AF allele frequency, MGRB Medical Genome Reference Bank, NUMT nuclear mitochondrial transcript, WES whole exome sequencing

### Cohort frequency of identified pathogenic variants

Nearly half of the distinct variants identified (14/34) occurred multiple times in this modest cohort. Pathogenic variants at positions m.1555 (m.1555G>A; *n* = 5), m.3243 (*n* = 6; of which three were m.3243A>G and three were m.3243A>T) and m.5650 (m.5650G>A; *n* = 5) were most common. Additionally, m.14484T>C, m.1494C>T, m.11778G>A, m.14674T>C, m.616T>C and m.5703G>A variants were each found in three individuals, and m.1630A>G, m.3697G>A, m.9035T>C and m.10197G>A variants each occurred in two individuals. The remaining 20 variants all occurred only once in this cohort; as a result their population prevalence is likely overestimated due to the relatively small cohort denominator.

### Haplogroup background of identified pathogenic variants

The haplogroup structure of the cohort as outlined above comprised predominantly of haplogroups H (44.29%), U (13.69%), J (11.93%), T (10.80%) and K (8.17%). The proportion of individuals with pathogenic variants from the main haplogroups H (43.55%), U (19.35%) and J (9.68%) were consistent with their respective haplogroup proportions in the cohort. Pathogenic variation was relatively under-represented in haplogroup T (2 of 62, or 3.23% of individuals carrying pathogenic mtDNA variants, compared to 10.8% of cohort (odds ratio 0.28; 95% CI 0.03-1.05)), although this was not statistically significant (*p* = 0.06) (**Figure 2**). Minor haplogroups in aggregate were relatively but not significantly over-represented (OR 3.30, 95% CI 0.64-10.67, *p* = 0.07), with the wide confidence interval reflecting the small numbers per group. (**Figure 2**) Of note, homoplasmic pathogenic LHON m.14484T>C variants were identified in association with haplogroups H and U, whilst none occurred on the (high risk) haplogroup J background.[36]

**Figure 2.**
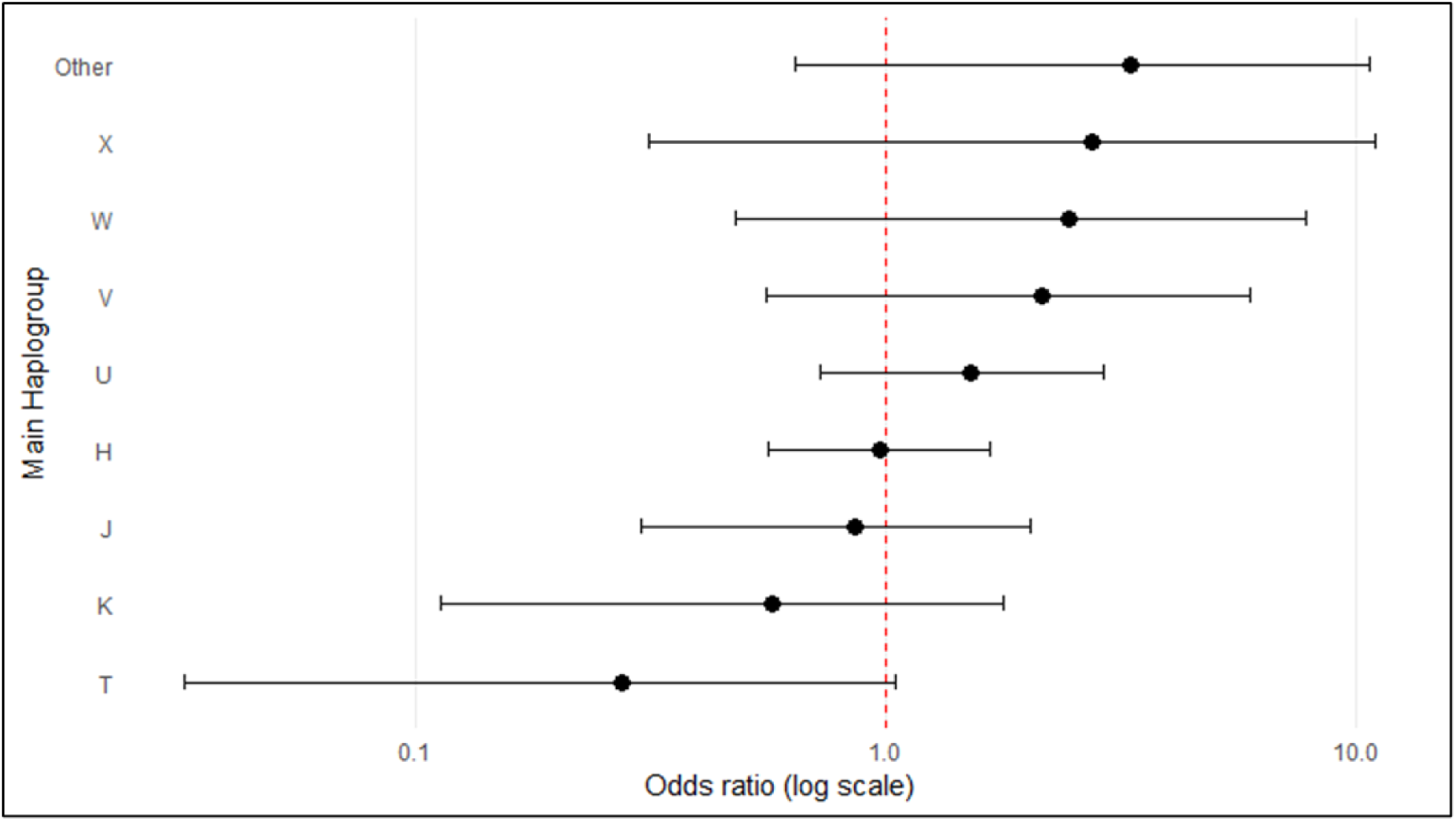
Haplogroup representation amongst individuals carrying pathogenic variants compared to haplogroup representation in the MGRB cohort, depicted as log odds ratio with 95% confidence intervals (CI), demonstrates no significant over- or underrepresentation of any haplogroups. “Other” contains haplogroups B, R and N, each with one pathogenic variant carrier.

### Heteroplasmy of identified pathogenic variants

Of the 62 pathogenic variants identified, 9 were found at or near homoplasmy (m.1494C>T (2), m.1555A>G (3), m.11778G>A (1), m.14484T>C (3)), whilst the remaining 53 variants occurred a variable heteroplasmy. Amongst these heteroplasmic variants, the mean heteroplasmy was 4.21% (median 1.9%, IQR 1.34-3.6%, range 1-36.63%). Of the 15 variants with heteroplasmy >10%, six were heteroplasmic variants (m.14674T>C (36.63%), m.9035T>C (20.84%), m.3255G>A (19.06%), m.8528T>C (13.9%), m.616T>C (11.53%), m.12201T>C (11.02%)), in addition to the nine homoplasmic variants described above. Considering a commonly implemented clinical heteroplasmy threshold of 5%, an additional four variants (m.9035T>C (8.37%), m.1494C>T (7.19%; although note hearing loss only with homoplasmy), m.3635G>A (5.98%; LHON only with homoplasmy) and m.7497G>A (4.97%)) would have been included. Of the 44 variants with heteroplasmy between 1 and 5%, mean heteroplasmy was 1.98% (median 1.66%, IQR 1.255-2.28%, range 1-4.97%).

### Recognised disease associations of identified pathogenic variants

Recognised disease associations for the homoplasmic variants identified are deafness (m.1555A>G and m.1494T>C) and LHON (m.14484T>C, m.11778G>A). These variants are well known to exhibit variable penetrance and disease risk, making their occurrence in a healthy population cohort unsurprising, albeit with implications for understanding penetrance.[36, 37] Amongst the identified heteroplasmic variants, the potential disease associations most commonly included myopathy / chronic progressive external ophthalmoplegia (CPEO; 35 of 62 variants, 1% of individuals in the MGRB cohort), movement disorders (29 variants, 0.83% of total cohort; predominantly ataxia and dystonia), cognitive dysfunction (28 variants; 0.8% of cohort), hearing impairment (27 variants; 0.77% of cohort) and seizures (24 variants, 0.69% of cohort; including myoclonus phenotypes). This highlighted a potential predominance of neurological system, vision and hearing involvement. Variants associated with endocrinopathy (23 variants; 0.66% of cohort) and cardiac dysfunction (21 variants; 0.6% of cohort) were also relatively common. Recognised mitochondrial syndromes that may be associated with the variants identified include Leigh Syndrome (12 variants; 0.34% of cohort), Mitochondrial Encephalopathy with Lactic Acidosis and Stroke-like episodes (MELAS; 16 variants; 0.45% of cohort), Myoclonic Epilepsy with Ragged Red Fibres (MERRF; 6 variants; 0.17% of cohort) and Mitochondrial NeuroGastroIntestinal Encephalopathy (MNGIE; 3 variants; 0.085%).

### Evaluation of polymorphic NUMT contamination

Evaluating the MGRB *mity* dataset (including the hypervariable region and considering Tier 1 and 2 *mity* variants; 363,250 variants) for the specified 25 NUMT-FP sites indicated that 61,187 variants (16.84% of total variants) mapped to one of these locations. This decreased to 57,392 (15.80% of total variants) when NUMT-FP variants were restricted to lower heteroplasmy (<10%) at which NUMTs might reasonably be expected to be observed. Evaluating variants with heteroplasmy ≥1%, the number of lower heteroplasmy NUMT-FP variants approximately halved to 33,085 (33,085 of 159,847 variants; 18.32% of variants with heteroplasmy ≥1%). Excluding the control region (recommended with *mity* analysis [43]), further reduces NUMT-FP variants to approximately 8.58% of total variants (8,011 of 93,316 variants with heteroplasmy ≥1%; although 19.85% of heteroplasmic variants 1-95%). However, as noted, numtB originates from the control region; excluding this region therefore disproportionately reduces apparent NUMT-FP variant contamination. Conversely, inclusion may overestimate NUMT FP variants (due to reduced coverage and consequent reduced reliability of sequencing in this region with *mity*). The proportion of low heteroplasmy variation attributable to NUMT-FP variants was higher at lower mtCN, and the heteroplasmy at which NUMT-FP variation was observed also varied by mtCN, although the relationship between mean heteroplasmies per sample and mtCN was observed to not be purely inverse (**Figure 3**).

**Figure 3.**
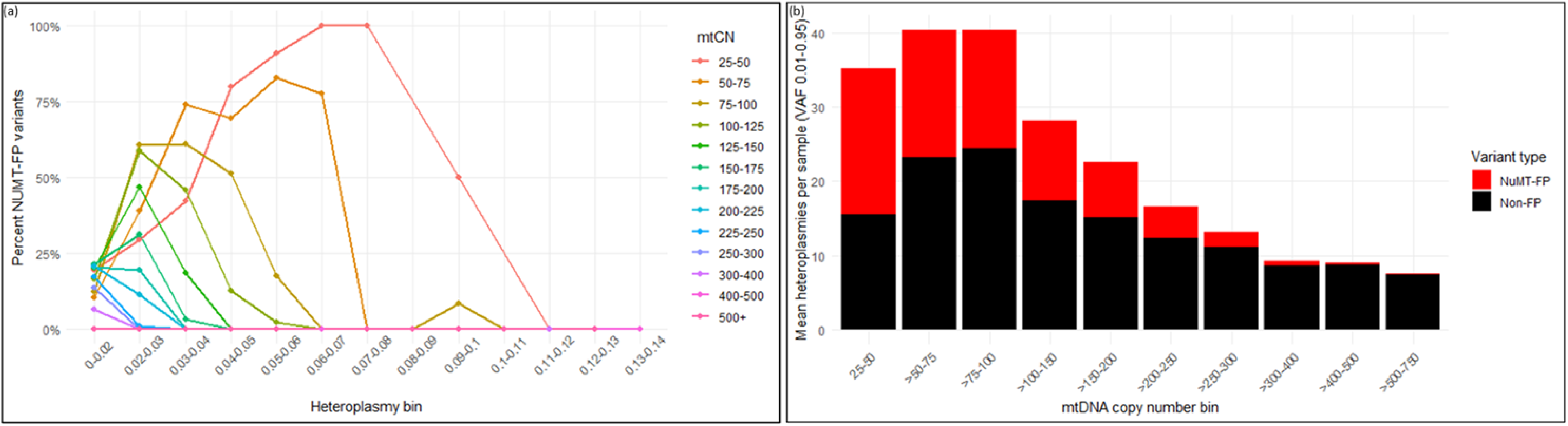
Relationship between mtCN, heteroplasmy and variation, particularly NUMT-FP variation (a) the proportion of NUMT-FP variants of total variants within heteroplasmy bins, stratified by mtCN, showing an increased proportion of NUMT-FP variants amongst individuals with lower mtCN, especially at very low heteroplasmy. Note that due to few samples with very low mtCN, the very high proportions above heteroplasmy 5% represent only 1 or 2 total variants. (b) relationship between mean heteroplasmies per sample and mtCN, with higher proportion of NUMT-FPs in samples with lower mtCN. The relationship between heteroplasmies and mtCN is also observed not to be purely inverse in this cohort.

Taking these NUMT-FP variant observations into consideration, we calculated the theoretic heterozygous NUMT-FP VAF (i.e., heteroplasmy) for each sample as 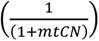 and compared this theoretic estimation with the observed heteroplasmy of identified NUMT-FP variants (**Figure 4**). The resulting distribution indicated two clusters, approximating *y* = *x* and *y* = 2*x*, suggesting the presence of heterozygous and homozygous NUMTs. To assess this, a two-component finite mixture model, fitted to the relationship between observed and expected heteroplasmy and constrained to pass through the origin, estimated slopes at 1.14 and 2.15 (**Figure 4**), closely approximating the theoretical expectation for heterozygous and homozygous NUMT carriers, respectively. There was also evident broadening of variance as theoretic NUMT VAF increased, corresponding with a decrease in mtCN. These observations indicated that a flat heteroplasmy threshold would differentially impact sample variant inclusion depending on mtCN (**Figure 4**), potentially missing a proportion of genuine mtDNA variants that occur at low heteroplasmy as mtCN increases and retaining NUMT-FP variants at higher apparent heteroplasmy as mtCN decreases. This suggested that a sample-specific heteroplasmy threshold based on theoretical NUMT VAF may better mitigate NUMT-FP contamination, enabling greater sensitivity for genuine low heteroplasmy pathogenic variant calls, whilst mitigating loss of non-NUMT-FP variant calls.

**Figure 4.**
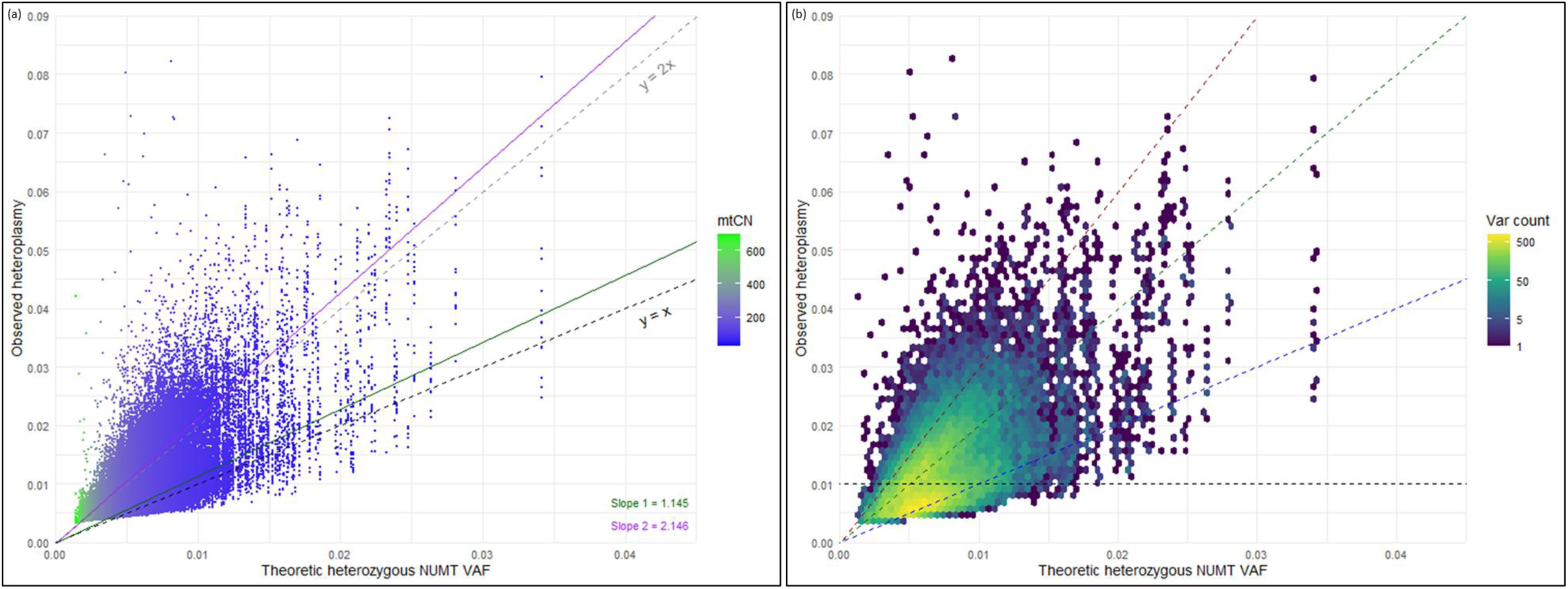
Theoretic and observed nuclear mitochondrial transcript-false positive (NUMT-FP) variant allele fraction (VAF; or effective heteroplasmy) **(a)** scatter plot of all NUMT-FP variants (heteroplasmy <0.1 or 10%) demonstrates two clusters with derived slopes (green and purple lines) approximating theoretic heterozygous (*y* = *x*) and homozygous (*y* = 2*x*, depicted by black and grey dashed lines, respectively) NUMT-FP VAFs. NUMT-FPs occur at higher apparent heteroplasmy with lower mitochondrial DNA copy number (mtCN). Variance of observed vs expected heteroplasmy is observed to increase with increasing theoretic VAF, corresponding with decreasing mtCN. The hex-binned density plot in **(b)** illustrates the ridges approximating *y* = *x* and *y* = 2*x*. The red, green and blue dashed slope lines indicate *y* = 3*x*, *y* = 2*x* and *y* = *x* respectively, demonstrating the number of NUMT-FPs (var count) that would be removed by using sample-specific VAF-scaled heteroplasmy thresholding, whilst the horizontal dashed line indicates effect on NUMT-FPs of a flat heteroplasmy threshold of 1% (VAF 0.01).

Implementing a sample-specific heteroplasmy threshold at twice or three times the theoretic NUMT VAF reduced NUMT-FP variant observations by two thirds (67.53%) and nearly 95% (94.27%) respectively (**Figure 5**). This was countered by only a modest reduction in pathogenic variant detection (indexed to those pathogenic variants identified with *mity* default ≥1% heteroplasmy threshold), to 79% and 61% respectively, whilst retaining a greater proportion of total identified variation (**Figure 5**). The effect of NUMT VAF-scaled heteroplasmy thresholding on the cumulative distribution of variants and pathogenic variants by heteroplasmy is shown in **Figure 5**. By comparison, implementing a flat heteroplasmy threshold (at 3 or 5%) to achieve an equivalent reduction in NUMT-FP substantially reduced the proportion of total variation retained and resulted in a greater reduction in sensitivity for pathogenic variant detection (to 41 and 29%, respectively). An evaluation of heteroplasmy threshold optimisation, delineating trade-offs between sensitivity for pathogenic variation, mitigation of NUMT-FP calls and retention of total variation, is depicted in **Figure 5**.

**Figure 5.**
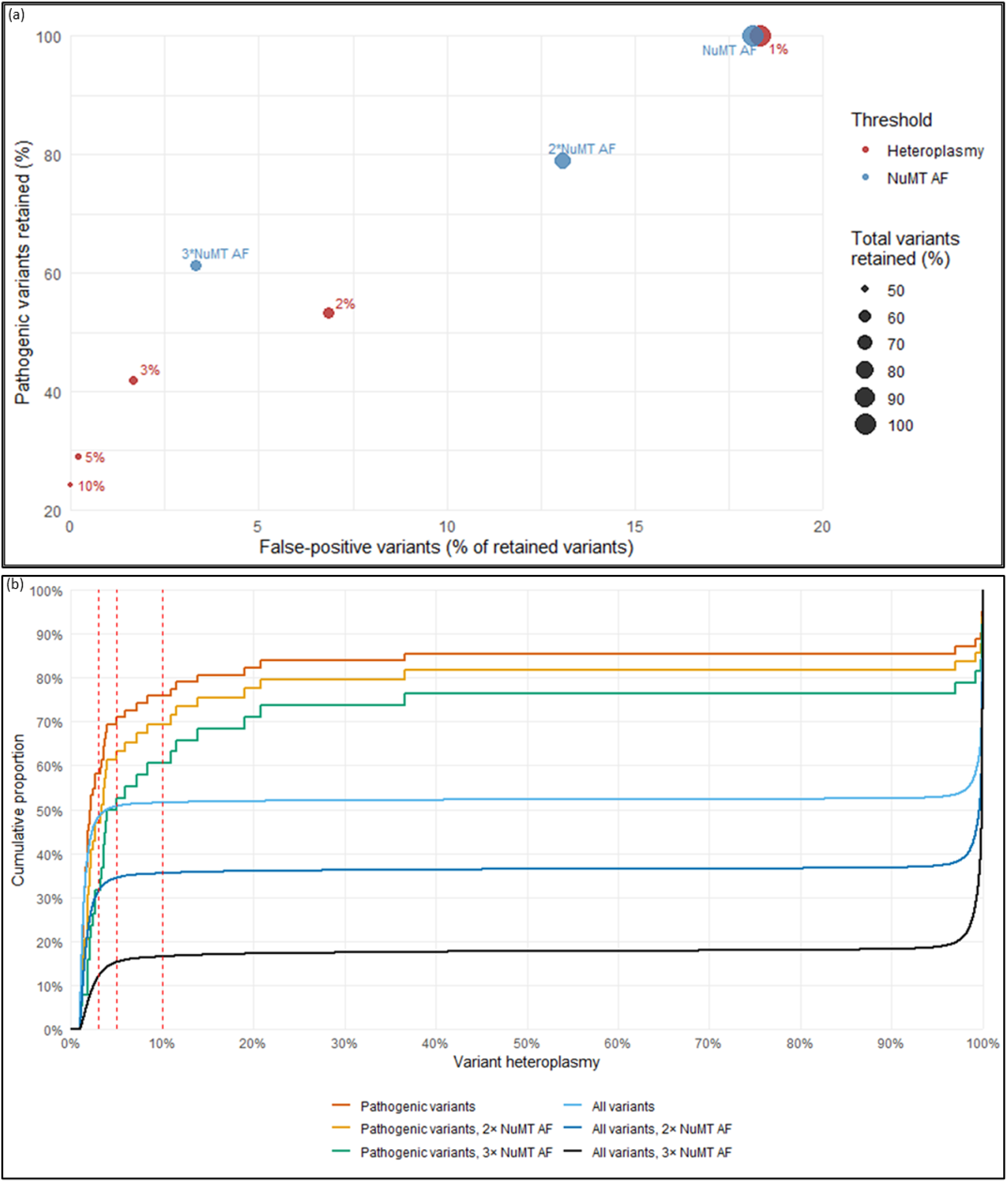
Effect of sample-specific nuclear mitochondrial transcript-allele fraction (NUMT-AF) scaled heteroplasmy thresholds, demonstrating **(a)** threshold optimisation and trade-offs, depicting the proportion of total variants retained (dot size), pathogenic variants retained (*y*-axis) and false positive variants remaining (*x*-axis, as a percent of total variants) with different heteroplasmy thresholding approaches (flat heteroplasmy thresholds in red compared to sample-specific NUMT-AF scaled thresholds in blue), and **(b)** effect on cumulative distribution of variants by heteroplasmy, indicating reduction in lower heteroplasmy variation with increasing thresholds. The red vertical dashed lines show the variation that would be excluded with fixed flat heteroplasmy thresholds at 3, 5 and 10% respectively.

### Comparison with publicly available genetic cohorts

To contextualise the pathogenic MD variant identification in the MGRB, we compared our findings to allele frequencies from large genomic database including gnomAD,[27] the UK Biobank[29] and HelixMTdb,[28] (**Table 2**) as well as estimates from prior disease and population-based cohort estimates (**Table 1**). Homoplasmic variant frequency was observed to be relatively consistent across estimates, whilst heteroplasmic variant frequency was more variable. To characterise the effect of heteroplasmy thresholds, the summed allele frequencies for all MitoMap [47] confirmed pathogenic SNVs from each database were compared. Implementing the sample-specific heteroplasmy thresholds in the MGRB at two and three times the theoretic NUMT VAF, as well as a flat 10% heteroplasmy threshold, we compared this to gnomAD with and without implementation of the low heteroplasmy filter (i.e., 10% and 1% heteroplasmy threshold) (**Table 2**). This clearly demonstrated the impact of heteroplasmy threshold selection: estimation of population frequency of pathogenic MD variants was more than halved with a 10% heteroplasmy threshold, compared to a 1% threshold. Implementation of sample-specific heteroplasmy thresholds set at twice and three times the theoretic NUMT VAF in the MGRB aligned summed allele frequency estimates from the MGRB very closely with those from gnomAD when implementing a 1% threshold. Despite some variability between population-based studies, clustering of population-based estimates appeared distinct from disease cohort estimates.

### Mitochondrial constraint measures

SCS ranged from 0.008 to 27.334 for individuals in this healthy older cohort, with a mean SCS of 4.362 (standard deviation 3.196). MCS ranged from 0.004 to 0.670, with a mean of 0.243 and standard deviation 0.089 (**Supplementary Figure 4)**. Mean constraint was slightly, although significantly (*p* = 0.002), higher in individuals carrying a pathogenic variant, compared to non-carriers (mean difference 0.038; 95% CI 0.014-0.06), whereas summed constraint was not significantly different (*p* = 0.162) between these two groups (mean difference 0.518; 95% CI −0.2136975 to 1.2493426). However, SCS was observed to correlate closely with the number of variants (r = 0.904, *p* <0.001) and increased slightly with advancing age (r = 0.125, *p* <0.001). This appears to be consistent with the observation of increasing mtDNA variation with age, and may reflect accumulation of somatic variation. MCS did not show significant association with number of variants (r = 0.011, *p* = 0.522) or the examined age span (r = 0.007, *p* = 0.678) (**Supplementary Figure 4**). An expected depletion of high heteroplasmy-highly constrained variants was observed (**Supplementary Figure 5**). This impact was notably weighted by the underlying variant heteroplasmy distribution, with greater representation of very low and very high heteroplasmy variants.

## Discussion

Recent advances in sequencing technology have enabled sensitive capture of mtDNA variation alongside nuclear DNA, facilitating comprehensive estimates of pathogenic mtDNA variation in population cohorts. In parallel, bioinformatic tools for analysing mtDNA continue to evolve, particularly balancing sensitive heteroplasmy detection against the risk of nuclear mitochondrial DNA segment-derived false positives (NUMT-FPs). Consequently, both understanding of population-level mtDNA variation and translation of this knowledge into the clinic is also evolving.

Our findings suggest that pathogenic mtDNA variants are more common in the population than previously estimated, with up to 1 in 56 individuals harbouring a potentially pathogenic variant. Despite the MGRB representing a risk-depleted cohort,[40] and in contrast to relatively reduced autosomal variation,[53] pathogenic mtDNA variation was more frequent than in other publicly available genomic databases.[27–29] Whilst heteroplasmy likely accounts for most of the difference, other contributing factors may include the relatively small denominator of the MGRB and the advanced age of participants, which associates with accumulation of somatic mtDNA variation.[40, 41, 54, 55] This is particularly relevant given the predominance of low heteroplasmy variation, and is unlikely to carry equivalent clinical risk.

Differences between studies are strongly influenced by heteroplasmy thresholds. Large population databases such as gnomAD and UK Biobank have applied more conservative thresholds than the 1% used here. [27, 29] Implementing equivalent thresholds (10% for gnomAD and 3% for UK biobank) to the MGRB yields similar variant frequencies (**Table 2**). However, while conservative heteroplasmy thresholds reduce false positives, they also substantially decrease detected variation – including pathogenic variants – by up to 75%. This is particularly relevant given the bimodal distribution of heteroplasmy, with a relative paucity of variants in the intermediate heteroplasmy range. Incorporating low heteroplasmy (1-10%) variants in the gnomAD database yields variant frequencies similar to those observed in the MGRB, while UK biobank estimates (heteroplasmy ≥3%) [29] fall between estimates with heteroplasmy thresholds set at 10% and 1%. Across datasets, population-level estimates of pathogenic mtDNA variation are more consistent with earlier population-based studies,[14, 15, 17, 18, 24] than with disease-cohort derived estimates,[21] suggesting that pathogenic variation may be more frequent, and variant penetrance substantially lower, than previously inferred.[36, 37, 53, 56] However, interpretation is further complicated by heteroplasmy, as variant effects are dose-dependent and influence both penetrance and clinical expression.[34, 57] Homoplasmic variant frequency estimates are relatively consistent across studies, whereas heteroplasmic estimates vary more widely. This likely reflects differences in sequencing pipelines, variant calling and heteroplasmy thresholds, as well as biological factors such as age, somatic variation, and population structure.

The pathogenic mtDNA variant frequency estimates here may therefore be considered an approximate “upper bound” for population variant prevalence. These estimates are relevant for interpreting clinical sequencing results and informing counselling, and underscore the need to integrate phenotypic data to better define penetrance and expressivity of variants. In this cohort, the predicted clinical phenotypes if variants were clinically manifest differed by variant type: vision and hearing loss for homoplasmic variants, whilst heteroplasmic variants were more frequently linked to neurological phenotypes, including myopathy, movement disorders, cognitive dysfunction and seizures. These patterns are consistent with reported healthcare utilisation in mitochondrial disease, with neurological system predominance.[25, 26] Although unlikely to be overtly expressed in this healthy, older population, these associations highlight priorities for large-scale phenotypic studies, in addition to those aspects considered by Cannon and colleagues.[29] Such work is essential to reconcile the gap between variant frequency and disease prevalence, and to determine whether reduced penetrance, mild or atypical expression, or sub-threshold heteroplasmy explains this discrepancy. This is particularly relevant given the limitations of blood heteroplasmy as a proxy for estimating heteroplasmy in other tissues.[35, 58] Improved understanding will have direct implications for clinical care, genetic counselling and reproductive decision making, especially as genomic testing becomes increasingly available.[59, 60]

Clinical interpretation of low heteroplasmy variation, particularly from blood samples, is inherently challenging. Conservative heteroplasmy thresholds are therefore commonly applied in clinical settings to reduce the risk of false-positive findings and misdiagnosis. However, such thresholds may exclude clinically relevant variants, given tissue-specific heteroplasmy and reports of disease at low blood heteroplasmy levels.[35, 57, 58] Accordingly, low level variants should not be dismissed outright, but instead require cautious interpretation supported by orthogonal validation. Characterising low heteroplasmy variation remains important for understanding the full spectrum of mtDNA variation and its contribution to disease risk. At a population level, this will refine estimates of disease burden and inform healthcare planning.[25] Continued advances in sequencing and bioinformatics, including long read approaches,[61] may help address challenges in distinguishing true mtDNA variation from NUMT artefacts, although rare and polymorphic NUMTs remain difficult to fully characterise.[30]

### NUMT contamination and heteroplasmy thresholding

Despite the development of tools such as *mity* to mitigate fixed NUMT misalignment, [43] the NUMT landscape remains complex due to ongoing integration of mtDNA into the nuclear genome.[30, 32, 52] Our findings support the presence of polymorphic NUMT-FPs, predominantly at low heteroplasmy, with contamination strongly influenced by heteroplasmy threshold selection and mtCN. Importantly, stringent exclusion of potential NUMT-FPs also removes a substantial proportion of true mtDNA variation, [27] differentially impacting samples depending on mtCN. We show that sample-specific thresholds, based on the theoretical heterozygous NUMT variant allele fraction, reduce NUMT contamination more effectively than fixed thresholds while preserving sensitivity for pathogenic variants. Although the optimal balance between sensitivity and specificity will depend on context, this approach offers a pragmatic strategy for improving mtDNA analysis. Future adoption of long read sequencing may ultimately resolve NUMT-related ambiguity at the point of sequencing.

### Mitochondrial constraint

Recent models of mitochondrial constraint provide a framework for evaluating tolerance to variation across the mitochondrial genome.[42] We applied mitochondrial local constraint (MLC) metrics to estimate cumulative variant burden (summed constraint) and average severity (mean constraint). Summed constraint strongly correlated with variant number and weakly with age, whereas mean constraint was increased in pathogenic variant carriers but stable across age and variant number, suggesting that accumulating mtDNA variation is not uniformly deleterious. This is consistent with evidence that much somatic variation is functionally neutral.[55] As expected, high heteroplasmy-high constraint variants were underrepresented. Extending these analyses to different age groups and disease cohorts may provide insight into mitochondrial contributions to ageing and neurodegeneration. However, further work – particularly incorporating heteroplasmy weighting and phenotypic data – is required to establish the clinical utility of constraint metrics.

### Limitations

The MGRB is relatively small, enriched for healthy older individuals and predominantly of European ancestry, [40] limiting generalisability. Additionally, mtDNA variation in blood is subject to negative selection [57, 62] and age-related somatic variation may influence the observed heteroplasmy spectrum. Despite this, comparison with external databases support the validity of our findings.[27, 29] Analyses of age-related variation suggest that the contribution of somatic mutations to overall variant burden is modest. Nevertheless, in clinical practice, low-level heteroplasmy – particularly in older individuals – requires careful interpretation and orthogonal confirmation.

Constraint analyses were limited by the lack of heteroplasmy weighting and potential residual NUMT contamination. Coverage limitations in the hypervariable region, due to alignment to the rCRS, may also affect some estimates, although alternative alignment strategies have mitigated this in other studies.[27] Acknowledging these limitations, we consider the characterisation of pathogenic variation across the heteroplasmy spectrum in these healthy older individuals a valuable addition to current knowledge, and the consideration of mitochondrial constraint measures as a potentially useful approach to quantifying mitochondrial quality.

## Conclusions

Sensitive analysis of mtDNA variation suggests that pathogenic variants may be present in up to 1 in 56 individuals. After accounting for heteroplasmy thresholds, these estimates are comparable across multiple population datasets and align with earlier population-based studies.

Inclusion of low heteroplasmy variants provides a more complete picture of population mtDNA variation, but introduces interpretive challenges in clinical settings. Conservative heteroplasmy thresholds reduce false positives and mitigate misdiagnosis risk, but at the cost of excluding potentially relevant variants. Accordingly, a balanced approach – incorporating thresholding, orthogonal validation, and clinical context – is essential.

Sample-specific heteroplasmy thresholds offer a promising strategy to maintain sensitivity whilst limiting NUMT-related artefacts. These findings may have important implications for genetic testing, counselling and healthcare planning, particularly as genomic sequencing becomes increasingly widespread.

## Supporting information

Supplementary File 1_NUMT FPs

## Declarations

### Funding

E.W was supported by a New Zealand Neurological Foundation Chapman Fellowship (1955CF).

P.L. is supported by a National Heart Foundation Future Leader Fellowship (grant no.: 107171) and National Health and Medical Research Council of Australia Investigator Grant (grant no.: 2026325)

R.L.D. was supported by an NSW Health Early-Mid Career Fellowship.

C.M.S was supported by a National Health and Medical Research Council Practitioner Fellowship (APP1136800)

### Conflicts of Interest

The authors have no relevant financial or non-financial conflicts of interest to disclose

### Ethics

The ASPREE Biobank study was approved by the Alfred Hospital Human Research Ethics Committee, and the study was performed in accordance with the ethical standards in the Declaration of Helsinki.

### Data Availability

The MGRB genetic data used for this study is available on request at https://sgc.garvan.org.au/terms/mgrb/index.html.

## Supplementary Materials

Supplementary File 1_NUMT-FPs Supplementary Figures

## Author Contribution

E.W. conceptualisation, data curation, formal analysis, funding acquisition, investigation, methodology, validation, visualisation, writing – original draft and review & editing

G.Q. data curation, formal analysis, writing – review and editing

S.R. data curation, resources and software, writing – review & editing

M.H. data curation, software, writing – review & editing

J.C. data curation, resources, software, writing – review & editing

C.Y. data curation, software, writing – review and editing

S.K. resources, software, writing – review & editing

C.L. conceptualisation, supervision, validation, visualisation, writing – review & editing

P.L. conceptualisation, investigation, resources, writing – review & editing

R.L.D. conceptualisation, methodology, formal analysis, administration, supervision, validation, visualisation, writing – review & editing

C.M.S. conceptualisation, funding acquisition, methodology, administration, resources, supervision, validation, writing – review & editing

## Use of AI statement

During preparation of this work, the authors used Google Gemma-3n-e4b AI Model on LM Studio to assist with editing. After using this tool, the authors reviewed and edited the content as needed, and take full responsibility for the content of the publication.

## Notes

### Competing Interest Statement

The authors have declared no competing interest.

