## Supplementary Figures for "Pathogenic mitochondrial genome variation, heteroplasmy thresholding and mitochondrial constraint measures in a healthy older cohort"

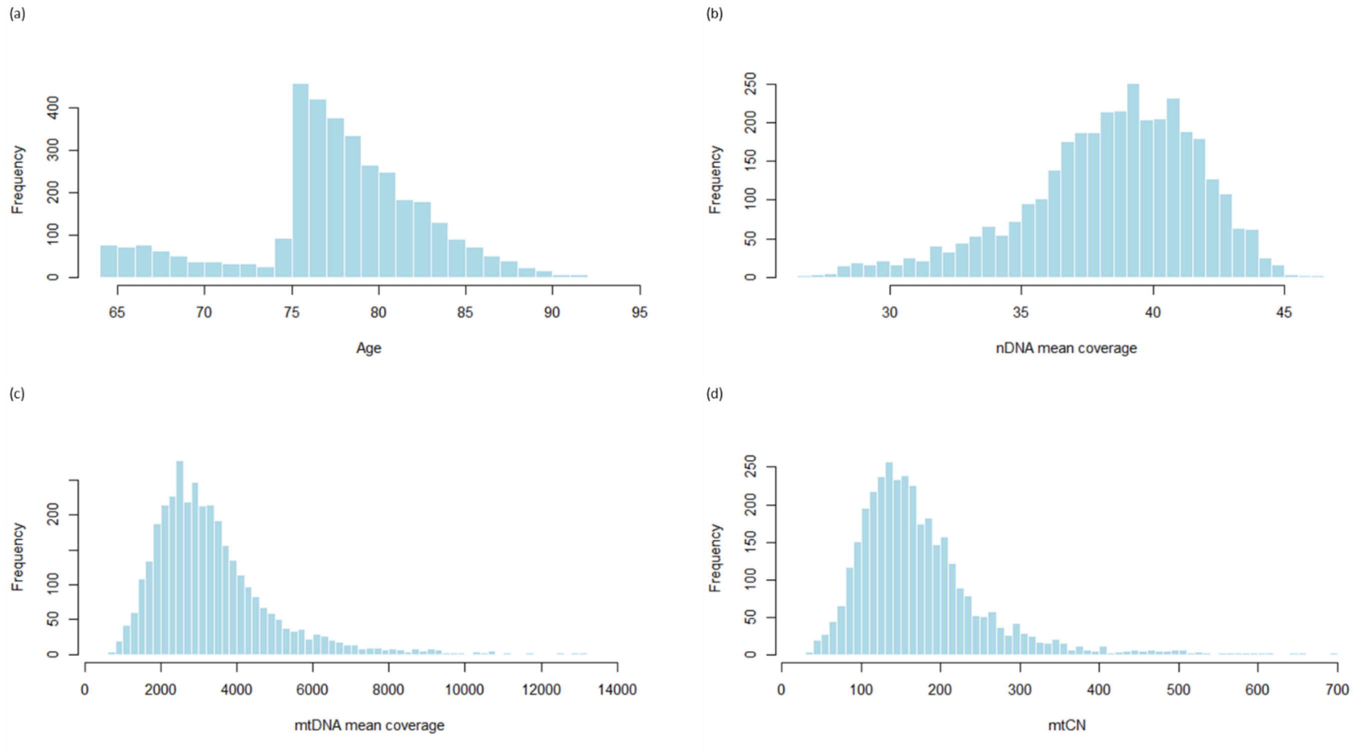


**Supplementary Figure 1** Distribution of participant ages **(a)** reflect a larger proportion of ASPREE (75+ years) samples in the cohort compared to 45 and Up samples. Mean coverage of nuclear DNA (nDNA) mean coverage centred around 30× coverage **(b)**, mean coverage of mitochondrial DNA (mtDNA) centred around 3300× **(c)** and mitochondrial copy number (mtCN) centred around 175 **(d)** in the MGRB cohort.

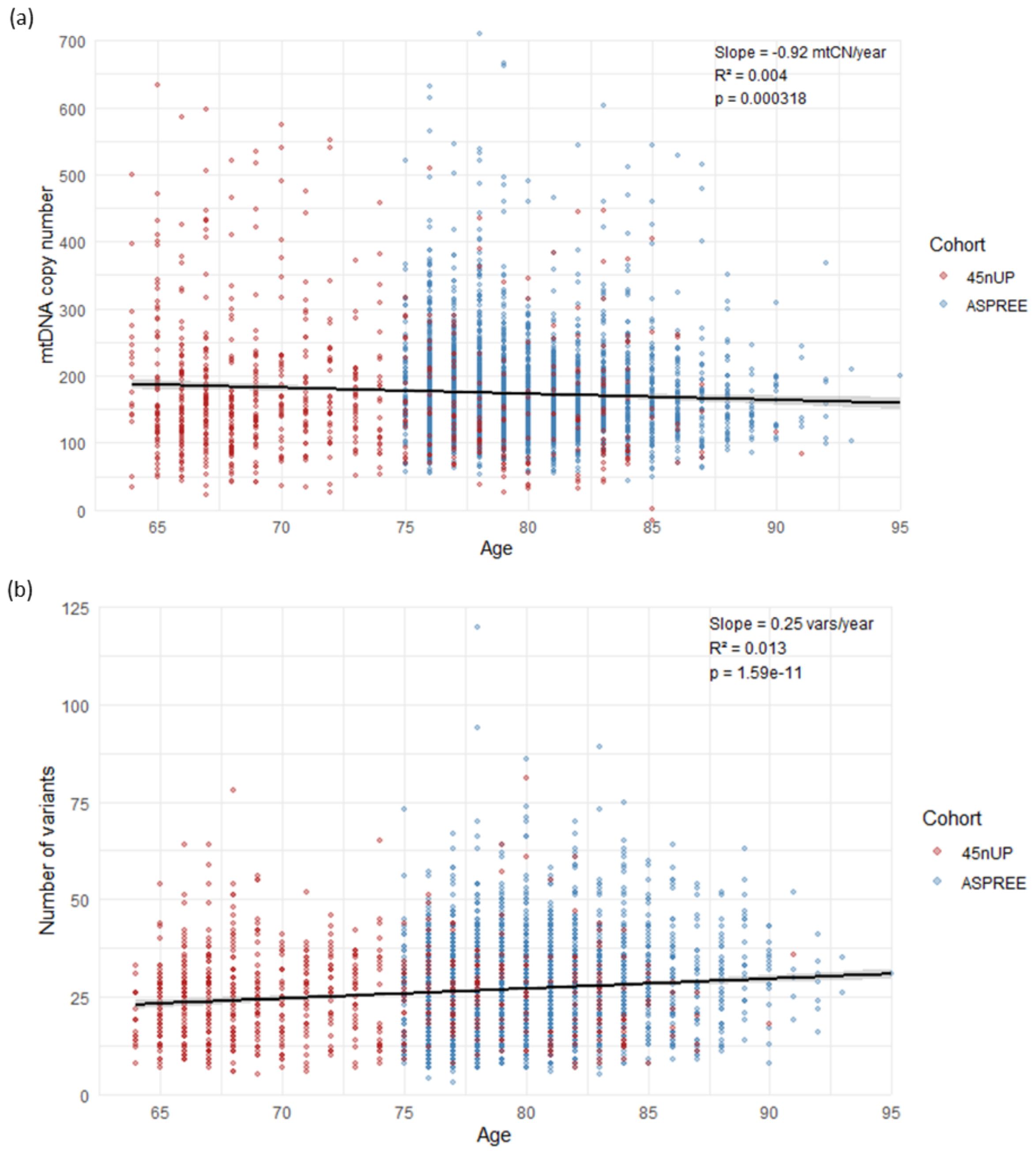


**Supplementary Figure 2** The relationship between age and mitochondrial copy number (mtCN) demonstrated an estimated decline of 0.94 copies per year **(a)** and the number of and number of variants (vars) per genome against age showed an estimated increase of 0.25 variants per year, or 2.5 per decade (b) across the 45 and Up and ASPREE study participants within the Medical Genome Reference Bank cohort.

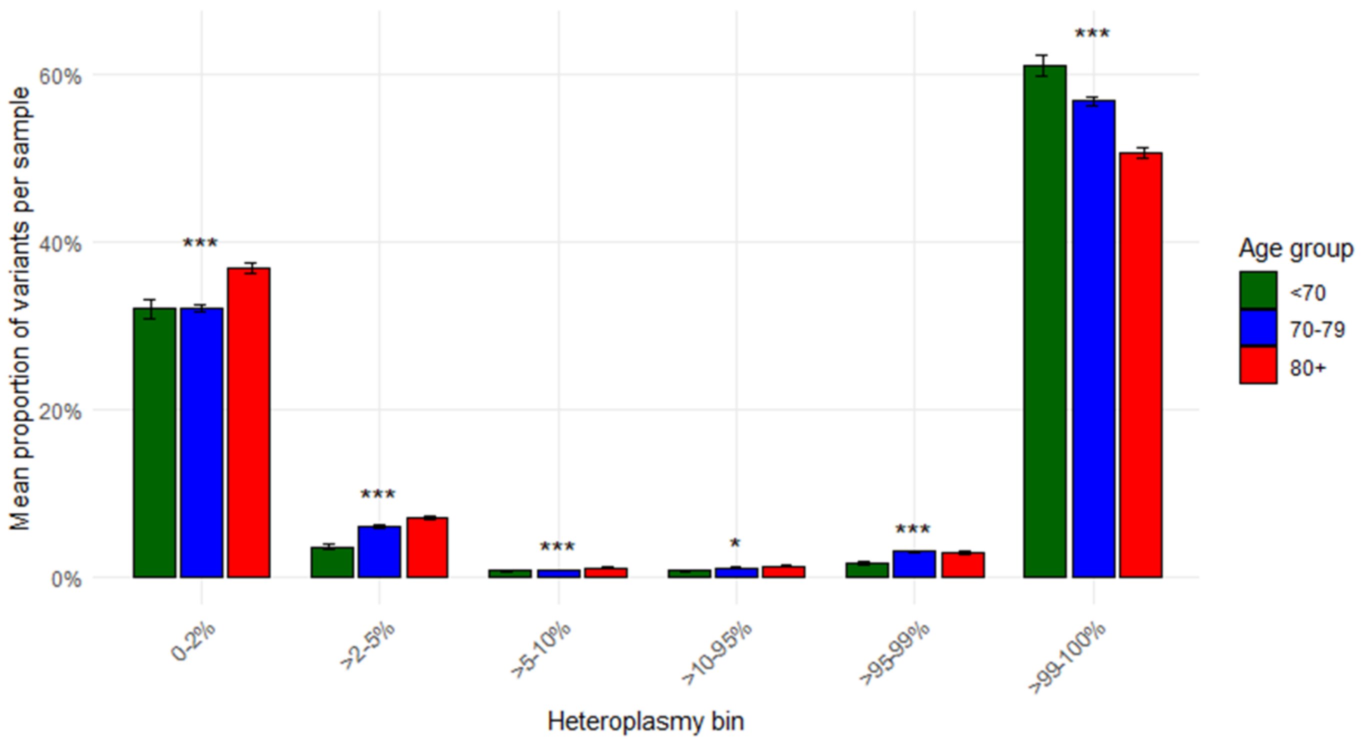


**Supplementary Figure 3** Proportion of variants per sample by age and heteroplasmy bin. Older individuals have a relatively greater proportion of low heteroplasmy variants and a correspondingly lower relative proportion of very high heteroplasmic and homoplasmic variants. * $p < 0.05$, *** $p < 0.001$.

Kruskal-Wallis test for difference in variant proportions between age groups at different heteroplasmy levels showed significant differences in all heteroplasmy groups (significance level denoted by stars * $p$ <0.05, *** $p$ <0.001).

Subsequent pairwise comparison at each heteroplasmy level indicated:

- significantly greater proportions of variants at very low heteroplasmy (0-2%) in individuals 80+ compared to <70 and 70-79 years
- at 2-5% heteroplasmy significantly greater proportions of variants in 70–79 year olds compared to those <70 years, and in 80+ year olds compared to both 70–79 year olds and <70 year olds.
- at 5-10% and 10-95% heteroplasmy, a significantly greater proportion of variants in 80+ year olds compared to 70-79 and <70 year olds.
- at high heteroplasmy >95-99%, the proportion of variants in 70-79 and 80+ year olds was significantly higher compared to <70 year olds
- at very high heteroplasmy and homoplasmy (>99-100%), a significantly lower proportion of variants in 70–79 year olds compared to <70, and in 80+ year old individuals compared to both 70-79 and <70 year olds.

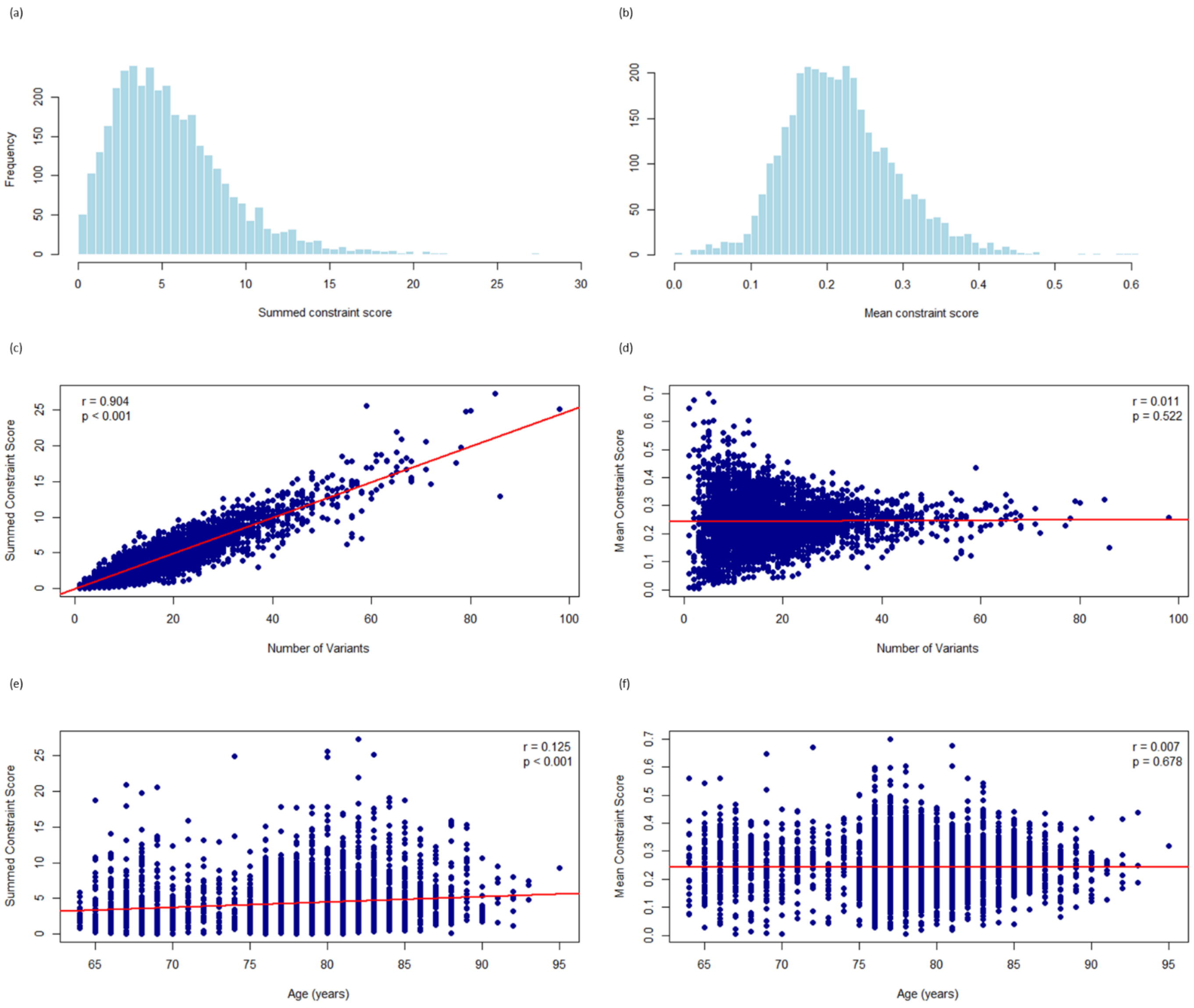


**Supplementary figure 4** Distribution and associations of summed and mean constraint scores for individuals in the medical genome reference bank (MGRB). Distribution of summed constraint scores (SCS) around a median of 3.6 **(a)** and of mean constraint scores (MCS) clustering around a mean of 0.24 **(b)** amongst individuals in the medical MGRB. Plotting SCS against number of variants for each individual in the cohort demonstrates an expected strong correlation between number of variants and SCS **(c),** whilst MCS does not increase with number of variants **(d).** SCS also shows a positive correlation with age **(e)** and MCS does not correlate with age **(f)**.

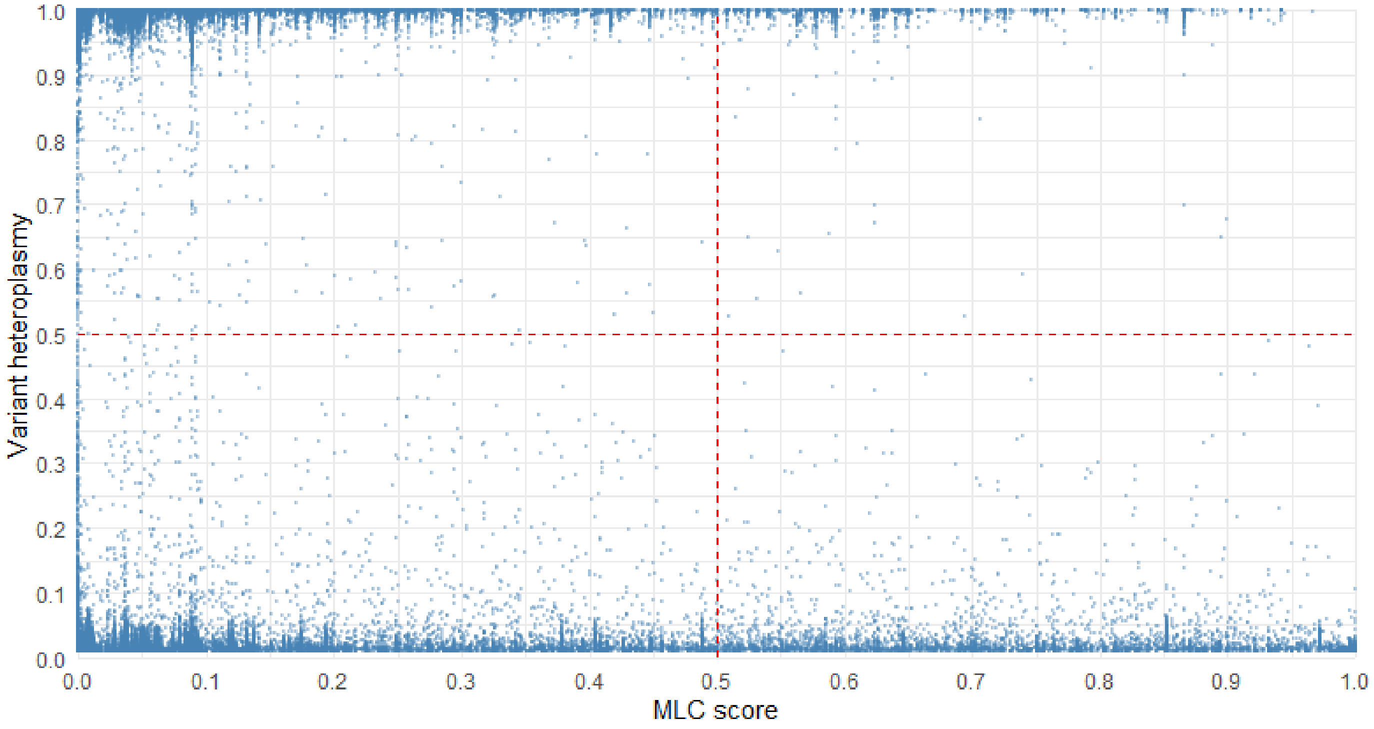


**Supplementary Figure 5** Plotting variant heteroplasmy against mitochondrial DNA local constraint (MLC) score, demonstrated an anticipated relative depletion of high heteroplasmy-high constraint variants (upper right quadrant) in this healthy elderly cohort. Conversely, there was a relative excess of low MLC score variation across the heteroplasmy spectrum (far left of upper and lower left quadrants).
